# Bias in the estimated reporting fraction due to vaccination in the time-series SIR model

**DOI:** 10.1101/2024.05.15.24307431

**Authors:** Tiffany Leung, Matthew Ferrari

## Abstract

The time-series Susceptible-Infectious-Recovered (TSIR) model has been a standard tool for studying the non-linear dynamics of acute, immunizing infectious diseases. The standard assumption of the TSIR model, that vaccination is equivalent to a reduction in the recruitment of susceptible individuals, or the birth rate, can lead to a bias in the estimate of the reporting fraction and of the total incidence. We show that this bias increases with the level of vaccination due to a double-counting of individuals who are infected prior to the age of vaccination. We present a simple correction for this bias by discounting the observed number of cases by the product of the number that occur prior to the average age of vaccination and the vaccination coverage during the initial susceptible reconstruction step of the TSIR model fitting. We generate a time series of measles cases using an age-structured SIR transmission model with vaccination after birth (at 9 months of age) and illustrate the bias with the standard TSIR fitting method. We then illustrate that our proposed correction eliminates the bias in the estimated reporting fraction and total incidence. We note further that this bias does not impact the estimates of the seasonality of transmission.

## 1 Introduction

The incidence of acute, immunizing infectious diseases displays complex non-linear dynamics that have been remarkably well described by simple dynamical models [1, 2, 3]. The time-series Susceptible-Infectious-Recovered (TSIR) model [2] has been an essential tool in bridging non-linear epidemic models and empirical data on disease incidence, particularly of immunizing infectious diseases during the pre-vaccination era [2, 4, 5, 6]. The TSIR model is a discrete time version of the continuous compartmental Susceptible-Infectious-Recovered (SIR) type epidemic models based on mass-action transmission [2].

The TSIR model makes an implicit assumption that the sum of (unimmunized) births and cases will be approximately equal over time. That is, each individual born will become infected (immunized) at some point over their lifetime and fast-scale deviations in the balance of cases and births reflect the non-linear dynamics of transmission and immunity, and seasonal variation [2]. For childhood infectious diseases, such as measles, mumps, and rubella, this is likely to be a fairly weak assumption and is supported by high adolescent seroprevalence in the pre-vaccine era [7]. A powerful property of the TSIR model, that derives from this assumption, is that it does not require incidence to be perfectly observed. The under-reporting fraction is encoded in the ratio of cumulative cases to cumulative unimmunized births [2, 6, 4].

Routine infant vaccination has been incorporated into the TSIR model by modulating the birth rate of unimmunized (susceptible) individuals. Births at each time step are discounted by the fraction immunized through vaccination after accounting for effectiveness [8, 9, 10, 11]. However, where vaccination takes place after birth, such as at 9 months of age for measles as recommended by the World Health Organization for countries where measles is common [12], a scenario arises in which an individual destined to be vaccinated may become infected in the interval following loss of maternal immunity and before vaccination age. Such an individual would be doubly counted in the TSIR model as both an immunized birth and a case, leading to the violation of the assumption that the sum of unimmunized births and cases equate each other over time.

In this study we show that this double counting in the TSIR model can lead to a bias in the estimation of reporting fractions and of disease burden in the presence of vaccination. We show that the magnitude of this bias increases with the coverage of routine vaccination and is stronger with higher birth rates. We also show that the bias does not impact the estimation of seasonality in transmission. We propose a method to correct this bias and evaluate its performance.

## 2 Methods

### 2.1 Dynamical model of measles

We generate daily time series of measles incidence using an age-structured stochastic compartmental model of measles transmission and vaccination, with a population stratified into 40 discrete age classes (24 monthly classes up to 2 years old; 1 class from 2 to 5 years old; 14 5-year classes from 5 to 75 years old; and 1 class of 75 years and older). Individuals in the model are either unvaccinated or vaccinated. Within each vaccination status, they may be temporarily immune with maternal antibodies (M), susceptible (S), infectious (I), or recovered (R).

Individuals are born either susceptible or temporarily immune with maternal antibodies. The fraction, *q*(*t*), of the birth cohort at time *t* that is born with maternal antibodies is determined by the proportion of individuals between 10 to 55 years old that is immune, either through prior infection or vaccination [13]. We assume a constant death rate for all age classes and set it equal to the birth rate for a constant population size. We used exponential rates of aging every 15 days. We assume maternal immunity to last an average of 4 months and lifelong immunity following infection or vaccination.

We use an age-dependent rate of routine vaccination, shaped as a truncated normal distribution with mean of 9 months and standard deviation of 0.5 months, constrained to the interval of 0 and 24 months. The height of the normal distribution is scaled by a constant factor such that the cumulative probability of vaccination by 24 months of age is equal to the routine vaccination coverage for a given simulation. We do not model a second dose of vaccination. Individuals who are maternally immune, susceptible, or recovered may become vaccinated. However, the vaccine is only effective for susceptible individuals. It has no effect on those who are maternally immune or who have recovered. We assume 100% vaccine effectiveness.

For simplicity, we present the model with the following set of ordinary differential equations:

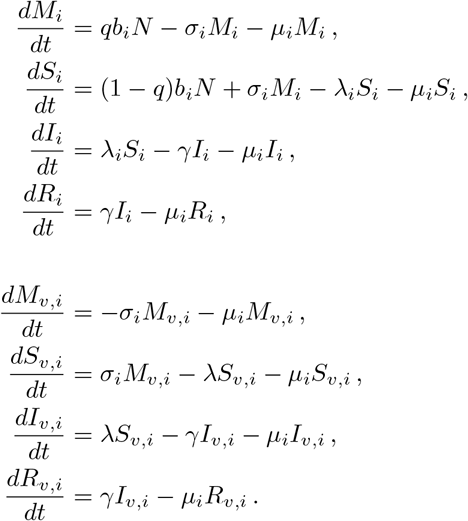

The force of infection for age class *i* is

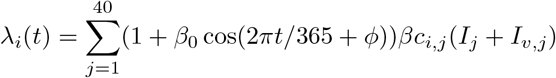

where *β* is the transmission coefficient; *c*_*i,j*_ is the contact rate from age group *j* to age group *i*; *β*_0_ is the relative amplitude of seasonal forcing; and *ϕ* is the phase shift. For the stochastic model and the full transition matrix, see the Supplementary Information.

The transmission coefficient is calculated with the next-generation matrix using a basic reproduction number, *R*_0_, of 15 [14, 15] and an infectious period of 14 days [2, 4]. Case importations occur with a probability 0.003 at all time steps to maintain endemicity. We assume either homogeneous contact mixing or assortative mixing following the pattern for Kenya [16].

We simulated the system in daily time steps using the tau-leaping algorithm [17], starting with 5 infected individuals in a fully susceptible population of 1 million with an age distribution consistent with Kenya [18]. The number of events is simulated as a Poisson draw with mean *k*_*i*_, where *k*_*i*_ is the rate of event *i* per time step. If the Poisson draw is higher than what is feasible, the number of events is set to the maximum allowable number.

We simulated 200 years of time series at a given vaccination coverage following a discarded initial transient period of 500 years (300 years without vaccination and 200 years with vaccination at the given coverage) to reach equilibrium. We considered vaccination coverage ranging 0–95% and multiple birth rates corresponding to that in Ethiopia and Chad (30 and 40 births per 1000 persons per year respectively [19]), amplitude of seasonal forcing (0.3 and 0.4), and phase shift (Day 0 and 180 of the year). For each setting, we divided the time series into 20 individual, non-overlapping 10-year time series and assumed that cases were observed at reporting fractions of 0.01, 0.05, 0.10, and 0.20 simulated as binomial draws from the true incidence at each time step. For each 10-year time series of observed cases, we fit the TSIR model (described in the next section) with and without correction for the double counted cases. We present the estimates of the reporting fraction, the reconstructed 10-year burden of measles, the estimated seasonal amplitude, and the estimated phase shift.

### 2.2 Fitting the TSIR model

The discrete TSIR model is described by the following equations [2, 4]:

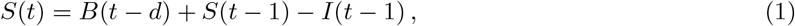

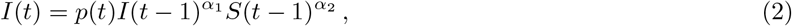

where *B*(*t* − *d*) represents births; *d* is the average duration of immunity derived from maternal antibodies; and *t* represents biweekly time steps (consistent with the infectious period for measles [2, 4]). The parameter *p*(*t*) is a time-varying transmission parameter with a 1-year (26 biweeks) period represented by

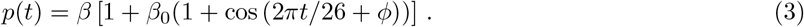

The coefficients *α*_1_ and *α*_2_ are cooperativity parameters, where *α*_1_ = *α*_2_ = 1 gives the standard mass-action transmission [20]. The parameters *β*_0_ and *ϕ* are the amplitude and phase shift respectively.

The estimated disease burden can be obtained by fitting *I*(*t*) of the TSIR model by first estimating the reporting fraction, *r*. We start with the simulated case data and estimate *I*(*t*) through its relation with the number of reported cases *C*(*t*) by

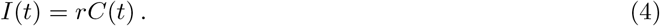

The reporting fraction, *r*, can be estimated using the linear regression relationship between cumulative cases and cumulative births, assuming births are known through available demographic data.

Unimmunized births, *B*(*t*), are calculated as a known constant birth rate multiplied by 1 − *V*, where *V* is the product of the fraction vaccinated and effectiveness, as used in other works [8, 11, 9]. Note that while we assume perfect effectiveness, these results are robust to any effectiveness as long as *V* is assumed known.

#### 2.2.1 Proposed correction to the double-counted cases

An individual may become infected between the time of loss of maternal immunity and vaccination and present as a case. The TSIR model would double count this individual as both an immunized birth and a case. We propose the following to correct these double-counted cases. When calculating the regression of cumulative cases against cumulative unimmunized births to estimate the reporting fraction, we subtract those cases that would be double counted. From each time step we subtract the cases that occur below the mean age of vaccination (9 months in our model), multiplied by the vaccination coverage (i.e., when coverage is low, fewer of these young cases would result in a double count). The adjusted time series of cases, *C*^*\**^(*t*), at age *a* is then

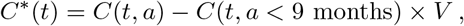

and the correction to Equation (4) becomes:

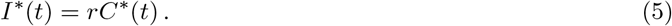

### 2.3 Fitting the amplitude and phase shift

We estimated the seasonality parameters (amplitude and phase shift) by fitting the the expanded transmission equation (2)–(3)

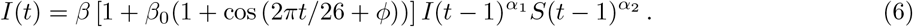

We begin by reconstructing *S*(*t* − 1) in Equation (1) by letting

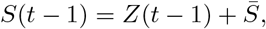

where *Z*(*t*) represents the residuals after regressing cumulative births and cumulative cases; and 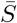 is inferred by maximizing the profile likelihood in a generalized linear model for Equation (2) without seasonality. Next we use a non-linear least squares to fit E quation (6) f or fi ve pa rameters: *α*_1_, *α*_2_, *β, β*_0_ an d *ϕ*. Th e first three parameters allow non-linear dynamics, and the latter two are our estimated seasonality parameters of interest: amplitude of seasonal forcing and phase shift.

Minimum bounds are set to (*α*_1_, *α*_2_, *β, β*_0_, *ϕ*) = (*ϵ, ϵ, ϵ*, 0, 0); the maximum bound for all parameters is ∞; and the initial starting point is (*α*_1_, *α*_2_, *β, β*_0_, *ϕ*) = (1, 1, *ϵ*, 0.5, 0).

In addition we estimate the reporting fractions under the scenario of increasing vaccination coverage (instead of a fixed coverage at equilibrium) with and without the proposed correction for double counted cases. We simulate 20 independent time series with increasing vaccination coverage from 60% to 90% over a 20-year period at an increasing coverage of 1.5% per year for birth rates ranging 10 to 80 births per thousand per year. This time series begins at 60% coverage, following a discarded initial transient period of 500 years (300 years without vaccination and 200 years with 60% vaccination coverage). For this we use only a single amplitude of 0.3 and phase shift of 0 days.

## 3 Results

For time series at equilibrium without vaccination, the estimated reporting fractions are unbiased for both birth rates and all combinations of true reporting fraction, amplitude, and phase shift, as expected (Figure 1A, G). At high vaccination coverage, however, there is a positive bias using the standard TSIR model fit for all combinations of reporting fractions, amplitude and phase shift (Figure 1 C–F, H–L). The bias is larger under a higher birth rate as a larger fraction of cases occur in young children below the age of vaccination (Figure 1). The biggest absolute error of the estimated reporting fractions using the standard TSIR model fit is observed at 9 0% vaccination coverage for a true reporting fraction of 0.20, estimated 0.278 and 0.246 for birth rates of 40 and 30 per 1000 respectively (Figure 1E, K, shown in red). This bias is also observed under an assortative (Kenya-like) contact mixing structure, under which violates the implicit assumption of the TSIR model (Figure S1). For example, at 60% vaccination coverage for a true reporting fraction of 0.10, the estimated reporting fractions are 0.154 and 0.117 under assortative and homogeneous contact mixing respectively (Figure S1).

**Figure 1:**
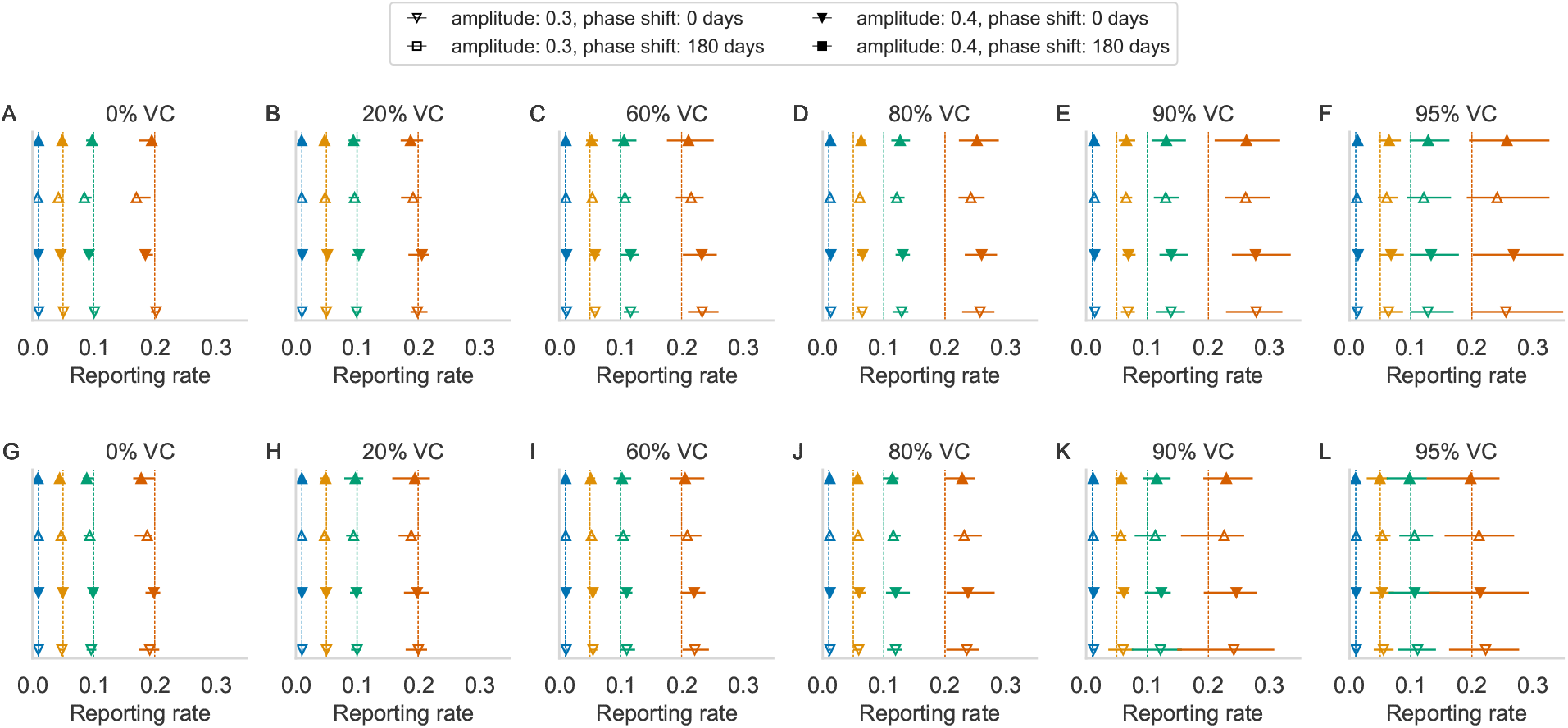
Estimated reporting fractions using the standard TSIR model for 40 (A–F) and 30 births (G–L) per 1000 persons per year and varying vaccination coverage (VC). Colors indicate different reporting fractions; vertical lines indicate simulated truth and symbols indicate mean and range (horizontal lines) of estimates from 20 simulations for different amplitudes (fill) and phase shifts (shape).

The estimated annual burden of measles is plotted against the true annual burden (Figure 2), forming an elongated shape along the line of unity to indicate years of low and high measles burden as in biennial dynamics. As expected, the total annual burden decreases as vaccination coverage increases. As a consequence of the positive bias in the reporting fractions, the estimated annual burden is negatively biased (Figure 2). This bias is observed for both birth rates and all chosen reporting fractions, amplitudes, and phase shifts. A regression of the estimated annual burden of measles against the true annual burden of measles shows underestimation at higher vaccination coverage, with the slope of the linear regression at a minimum of 0.77 at 90% and 95% coverage (Figure 2 E, F). At lower vaccination coverage, the burden is slightly overestimated from the truth, as observed by a slope of regression slightly above 1. This is due to a slightly underestimated reporting fraction at low vaccination coverage.

**Figure 2:**
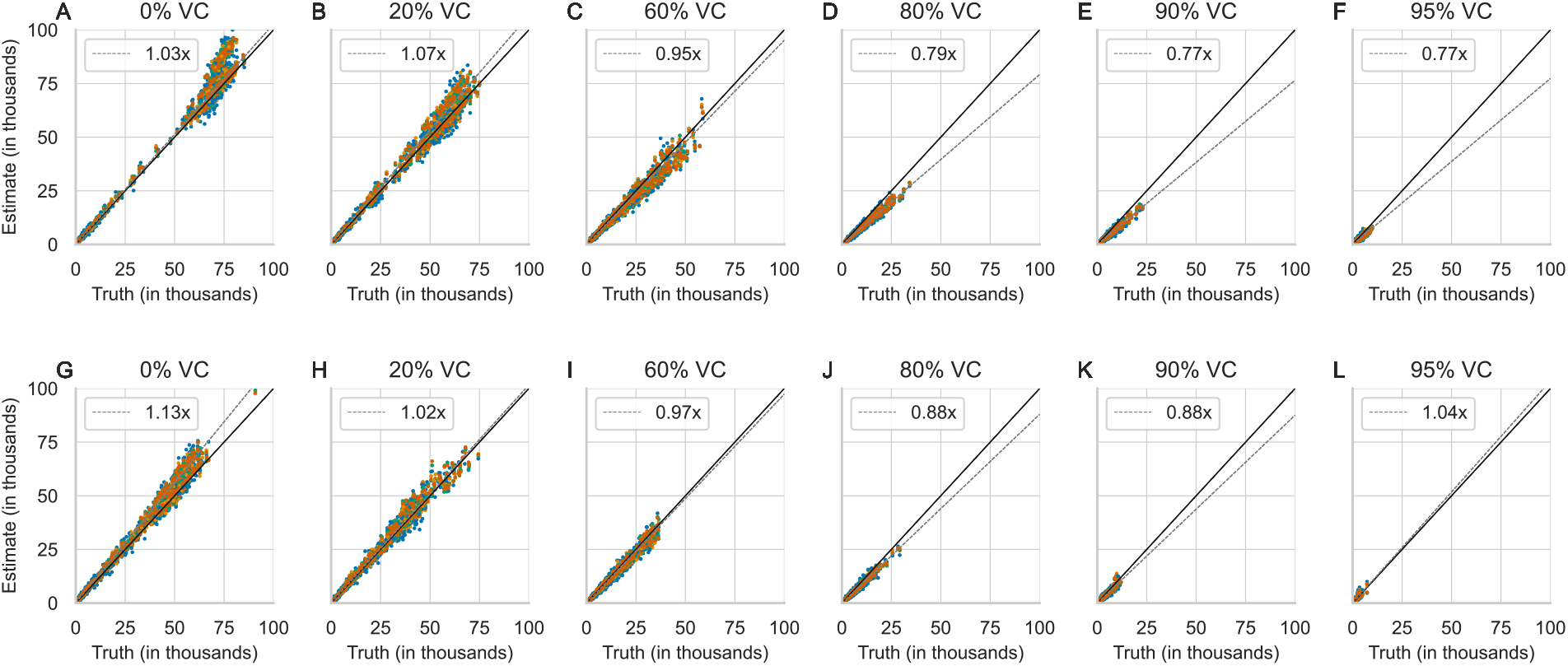
The estimated annual burden of measles using the standard TSIR estimator of the reporting fraction for 40 (A–F) and 30 (G–L) births per 1000 persons per year and vaccination coverage ranging 0– 95%. Diagonal lines are the line of unity (*y* = *x*; solid) and the regression of the estimated and true measles burden (dashed). Colors differentiate reporting fractions (0.01, 0.05, 0.10, and 0.20). Each reporting fraction shows 80 estimates (20 annual estimates per combination of amplitude and phase shift).

Correcting for the double-counting of cases occurring before the age of vaccination removes the positive bias in the estimate of reporting fraction (Figure 3). The biggest absolute difference of the estimated reporting fraction to true reporting fraction is now −0.03 with the correction (at 95% vaccination coverage at the higher birth rate; Figure 3F), compared to a maximum difference of 0.078 without correction. These reporting fraction estimates align more closely to the truth for both birth rates.

**Figure 3:**
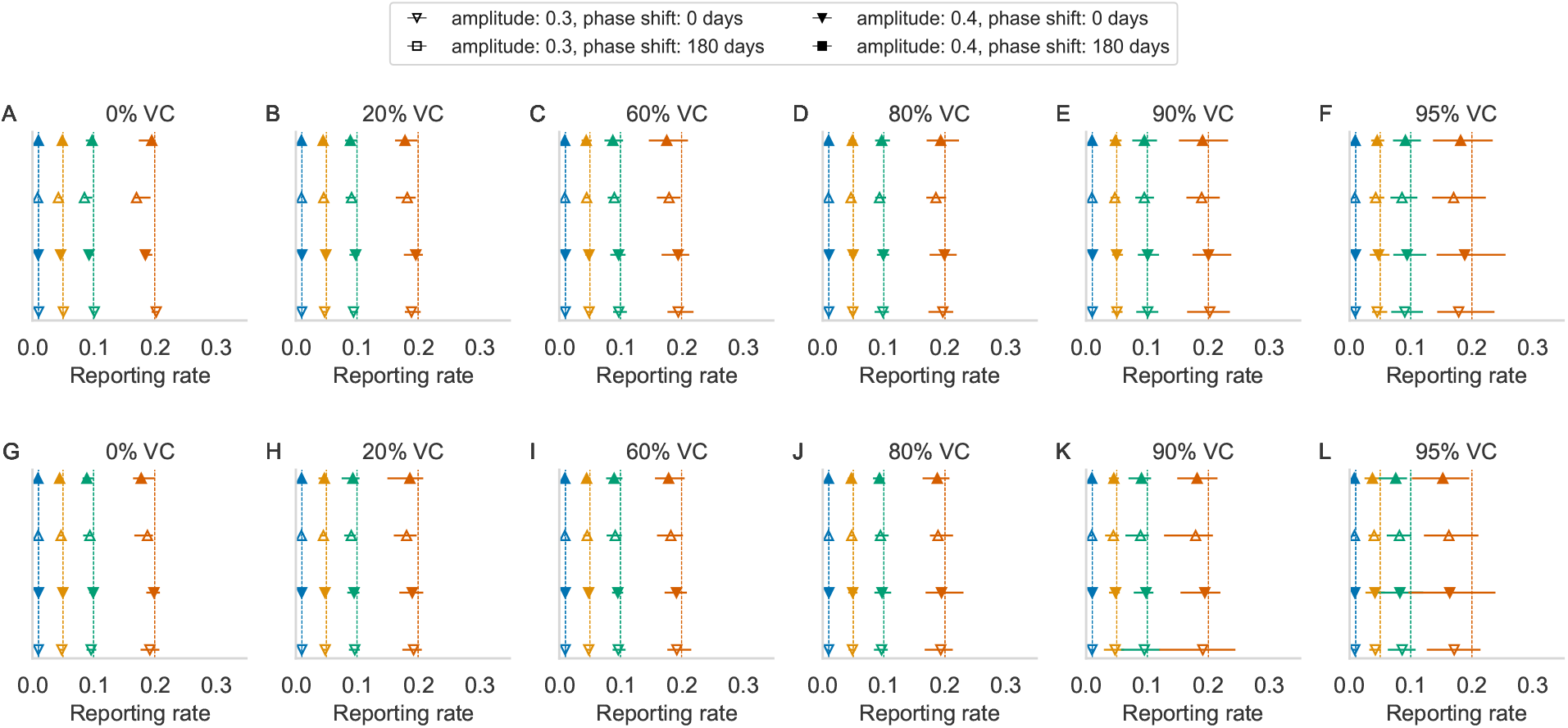
Estimated reporting fractions using the corrected TSIR estimator of the reporting fraction for 40 (A–F) and 30 (G–L) births per 1000 persons per year for vaccination coverage (VC) ranging 0–95%. Colors indicate different reporting fractions; vertical lines indicate simulated truth and symbols indicate mean and range (horizontal lines) of estimates from 20 simulations for different amplitudes (fill) and phase shifts (shape).

Using these estimates of reporting fraction with the proposed correction for the double-counting of cases also removes the negative bias in the estimated annual burden of measles (Figure 4). The regression of the estimated and true annual burden using the corrected TSIR estimator now shows an overestimation for higher vaccination coverage (Figure 4). For a birth rate of 40 per thousand per year, at 90% coverage with the corrected TSIR estimator, the estimated annual burden is now higher than the true annual burden by 5% on average (Figure 4E). This compares with the estimated burden using the standard TSIR estimator (without the correction), where at 90%, the estimated annual burden is smaller than the true annual burden by 23% on average (Figure 2E).

**Figure 4:**
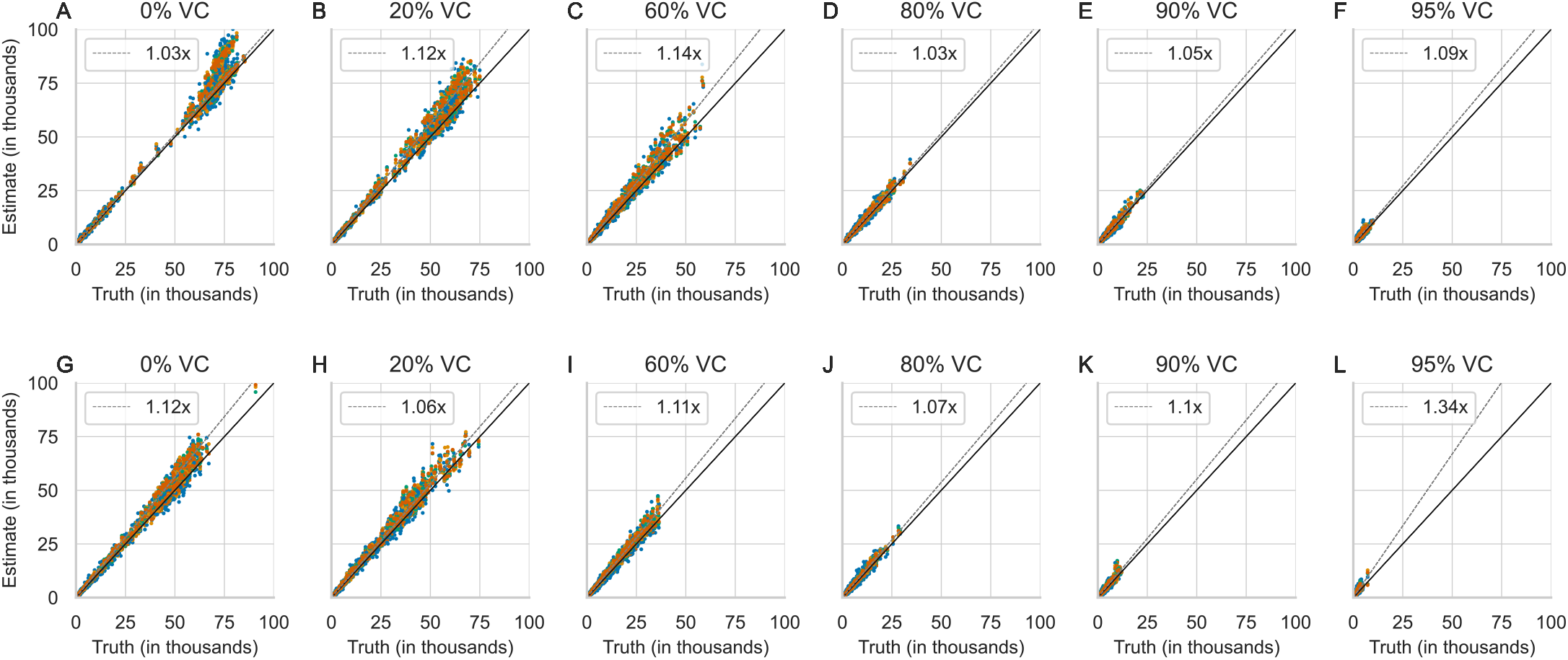
The estimated annual measles burden using the corrected TSIR estimator of the reporting fraction for 40 (A–F) and 30 (G–L) births per thousand persons per year and vaccination coverage ranging 0–95%. Diagonal lines are the line of unity (*y* = *x*; solid) and the regression of the estimated and true measles burden (dashed). Colors differentiate reporting fractions (0.01, 0.05, 0.10, and 0.20). Each reporting fraction shows 80 estimates (20 annual estimates per combination of amplitude and phase shift).

The estimated seasonality parameters are plotted against the true seasonality parameters using the standard TSIR estimator (without the proposed correction) in Figure 5 for the higher (A–L) and lower (M–X) birth rates. Despite the bias in the estimated reporting fraction and incidence, these estimates are unbiased for all 4 reporting fractions (Figure 5). Using the corrected TSIR estimator also does not affect the estimate of the amplitude and phase shift (Figure 6). However, the estimated amplitude is poor at 0.01 reporting fraction (blue) due to many zeros in the reconstructed incidence. The frequency of zeros make up as much as 60% of the reconstructed time series at 0.01 reporting fraction, compared with less than 10% of the same at 0.20 reporting fraction (Figure S2). A true reporting fraction of 0.05 and higher gives reasonably accurate estimated amplitude and range.

**Figure 5:**
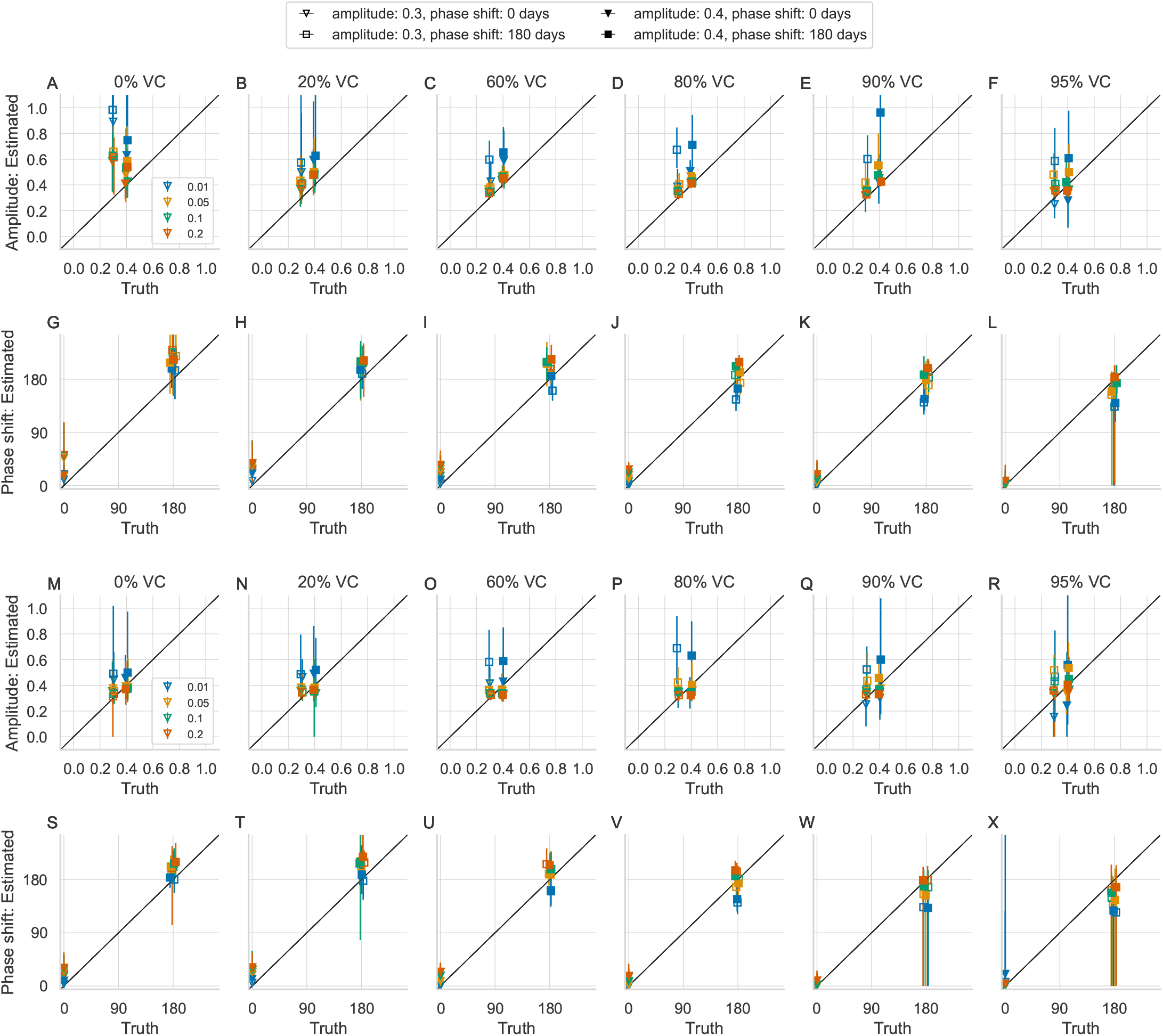
Estimated amplitude and phase shift using the standard TSIR estimator of the reporting fraction for 40 (A–L) and 30 (M–X) births per 1000 persons per year and vaccination coverage ranging 0–95%. Each panel shows the mean (marker) and range (vertical lines) over 20 ten-year intervals. Colors differentiate the 4 chosen reporting fractions.

**Figure 6:**
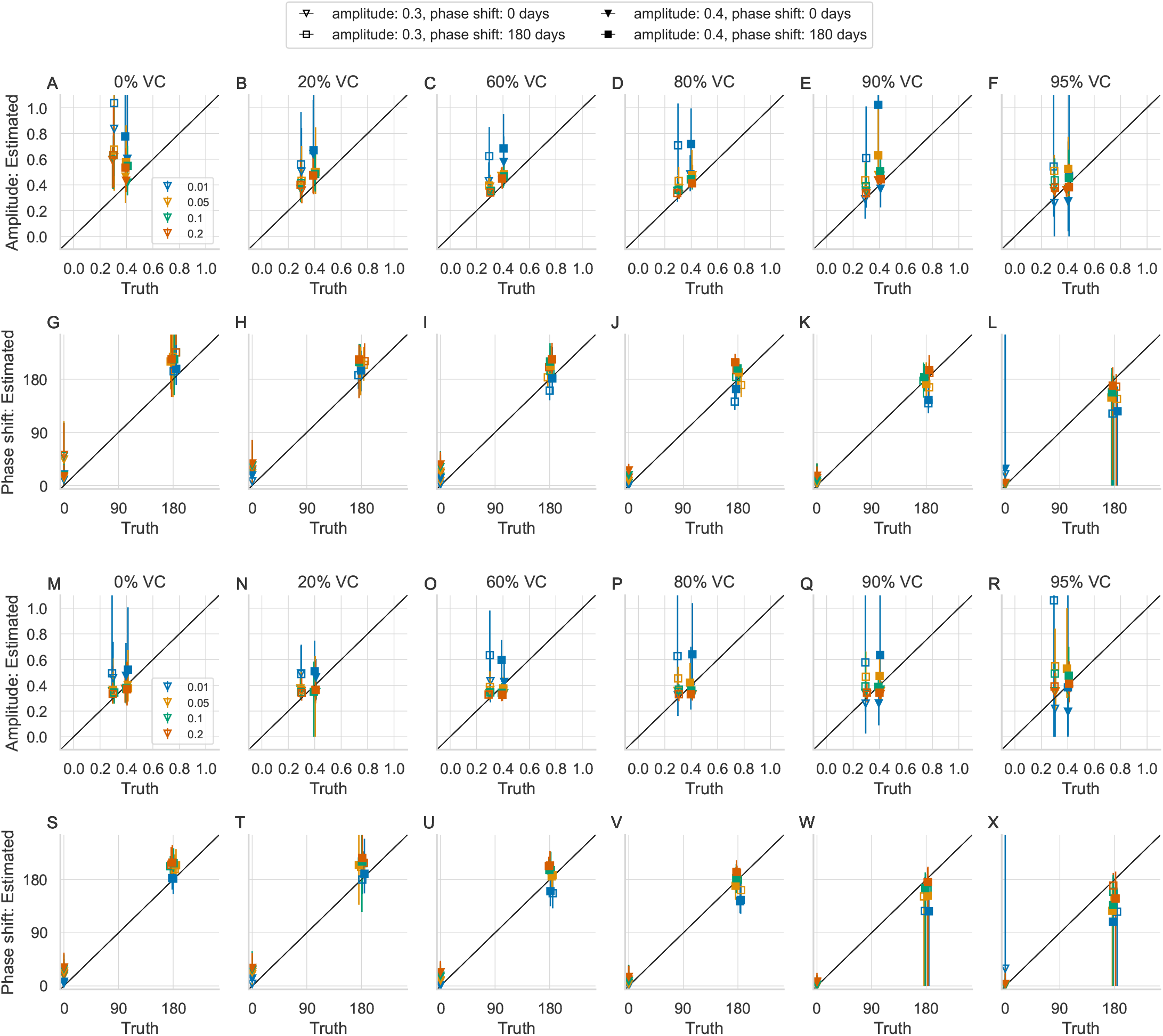
Estimated amplitude and phase shift using the corrected TSIR estimator of the reporting fraction for 40 (A–L) and 30 (M–X) births per 1000 persons per year and vaccination coverage ranging 0–95%. Each panel shows the mean (marker) and range (vertical lines) over 20 ten-year intervals. Colors differentiate the 4 chosen reporting fractions.

Realistically, few settings in the world have long-term equilibrium dynamics at a constant level of vaccination. The bias in the estimated reporting fraction that occurs at equilibrium is also present in transient time series dynamics and increases with birth rate (Figure 7). In the transient time series, vaccination coverage is increasing from 60% to 90% over 20 years (Figure 7). For a true reporting fraction of 0.20, the estimated reporting fraction without using the proposed bias correction is 0.211 at 10 births per 1000 and grows to an estimated 0.305 at 80 births per 1000 (Figure 7). Using the proposed bias correction overcomes this bias in the estimate of the reporting fraction for all birth rates and reporting fractions (Figure 7).

**Figure 7:**
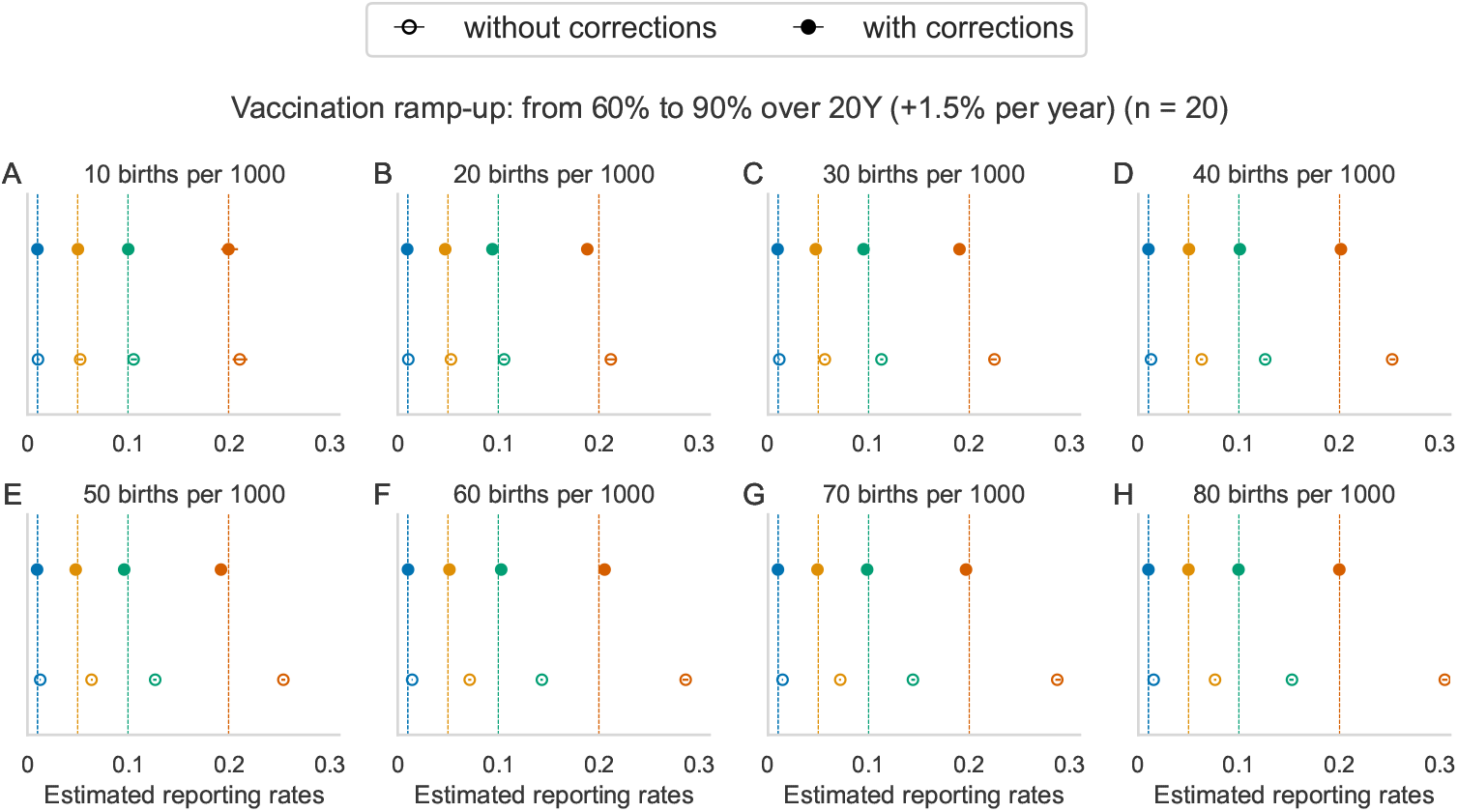
Estimated reporting fractions for simulations with increasing vaccination coverage from 60% to 90% over 20 years, at an increase of 1.5% coverage per year for reporting fractions (0.01, 0.05, 0.1 and 0.2; vertical lines) and different birth rates.

## 4 Discussion

We have illustrated that a standard simplifying assumption of the TSIR model, that the effect of vaccination can be represented as a reduction in the rate of susceptible recruitment, leads to a positive bias in the estimate of the reporting fraction and a negative bias in the estimate of the true incidence. When vaccination is not given immediately after birth, as is common for vaccine-preventable diseases such as measles, mumps, and rubella, then the standard assumption to the TSIR model to accommodate vaccination may lead to double-counting of young cases as immunized births. The magnitude of this bias will increase when birth rates are high (i.e., there are numerically many pre-vaccine age children to become infected) and when vaccination rates are high (i.e., a larger fraction of pre-vaccine age children who become infected would likely go on to be vaccinated). We proposed a method to correct for double counting and show that the correction led to a more accurate estimation of the reporting fraction and the incidence of infection. We also show that the estimation of seasonality parameters was not affected by the bias in the estimate of the reporting fraction. We note that the TSIR model was originally developed for application in a vaccine-free setting [2]. Thus, the bias we have identified is not a flaw of the model itself. Rather, it arises from the naive extension to settings with vaccination. Further, many applications of the TSIR model to fit observed time series have relied on time series that are not disaggregated by pre- and post-vaccine age individuals. The bias correction that we have proposed here requires the availability of age-specific case data, at least grouped as pre- and post-vaccine age. Where such data are not available, this analysis serves as a caution on interpreting the estimates of reporting fraction and incidence of infection in settings where the effect of the bias may be high.

The simplicity of the standard TSIR model and its ease of application for under-reported time series have made it an enduring tool for analysis of time series data even into the vaccine era [8, 11, 21, 10]. The bias identified here should be a cautionary note that simplifying assumptions, while often practically necessary, should be revisited as the quality and resolution of data increase.

## Data Availability

All data produced in the present study are available upon reasonable request to the authors.

## Supplementary Information

### Model description and equations

We used an age-structured discrete-time stochastic transmission model that incorporates both epidemiological and demographic transitions, building on a framework introduced by Klepac and Caswell [22] and Klepac et al. [23]. We describe the model structure following the notation of Metcalf et al. [24] and Winter et al. [25]. The model population is structured into an unvaccinated group and a vaccinated group. Individuals within each vaccination status group are further divided in epidemiological classes (temporarily immune with maternal antibodies, *M, M*_*v*_; susceptible, *S, S*_*v*_; infectious, *I, I*_*v*_; recovered *R, R*_*v*_) where the unvaccinated group is without subscript, and the vaccinated group is denoted by subscript *v*. The population is stratified into 40 discrete age classes (24 monthly classes from ages 0 to 2 years; 1 class from age 2 to 5 years; 14 5-year classes from age 5 to 75 years; and 1 class of age 75 years and older). At every time step a large transition matrix defines transitions from every possible epidemiological stage and age class combination to every other possible epidemiological stage and age class combination.

The transition matrix is described in two steps. First ignoring demography (aging and survival), we define matrix **A**_*a,t*_, which describes the epidemiological transitions within each age class *a* and discrete time-step *t* (set to 1 day), as

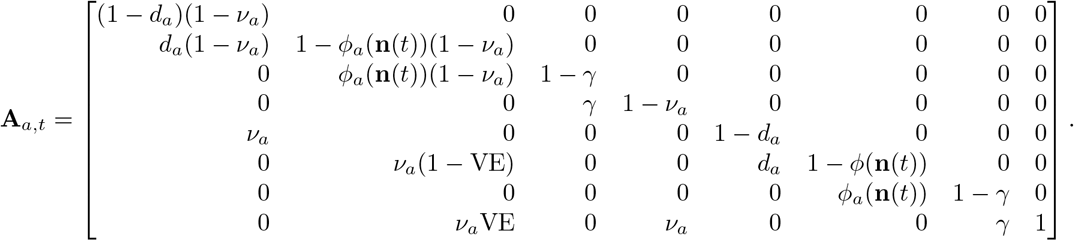

The eight rows and columns represent the stages *M, S, I, R, M*_*v*_, *S*_*v*_, *I*_*v*_, *R*_*v*_, respectively. In the transition matrix, *d*_*a*_ is the probability an individual in age class *a* loses maternal immunity; *ϕ*_*a*_ is the probability an individual in age class *a* becomes infected; *γ* is the probability of recovery; and *ν*_*a*_ is the probability an individual in age class *a* is vaccinated. A susceptible individual is successfully vaccinated with probability VE, reflecting the vaccine efficacy. Vaccination has no effect on individuals with maternal immunity or recovered individuals. The infection probability *ϕ* (also called the force of infection) is a function of **n**(*t*), a vector describing the population at time *t*

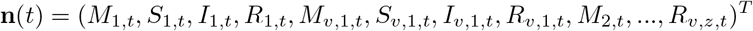

according to

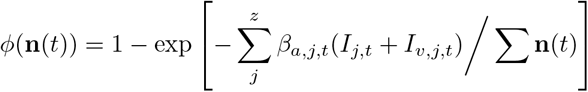

where *z* is the total number of age classes (*z* = 40 here); *β*_*a,j,t*_ is the transmission rate between individuals in age class *a* and *j* at time-step *t*; and *I*_*j,t*_ + *I*_*v,j,t*_ is the number of infected individuals in age class *j* at time-step *t*.

Transmission to individuals in age class *a* from individuals in age class *j* at time *t* is defined by *β*_*a,j,t*_ = *βc*_*a,j*_ (1 + *β*_0_ cos (2*πt/*365)), where *β* is the transmission coefficient calculated using the next-generation matrix; *c*_*i,j*_ is the mean contact rate between individuals in age class *j* to *i*; and *β*_0_ is the relative amplitude of seasonal fluctuations. The transmission coefficient was calculated with the next-generation matrix using a basic reproduction number *R*_0_ of 15 [14, 15] and an average infectious period of 14 days [2, 4].

Second, we define the full transition matrix, **A**(**n**(*t*)), that includes both epidemiological transitions from matrix **A**_*a,t*_ and demographic transitions (aging and survival). This matrix is used to project the entire population forward via aging, mortality, and infection dynamics according to:

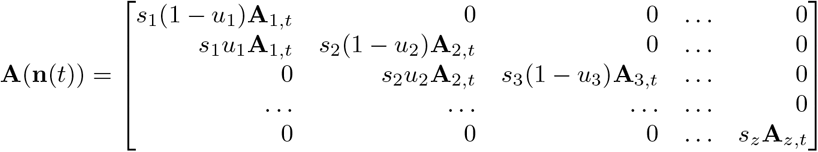

where *s*_*a*_ is the probability that an individual in age class *a* survives; *u*_*a*_ is the rate of aging out of age class *a*; and **A**_1,*t*_, **A**_2,*t*_, etc., are as defined in matrix **A**_*a,t*_. The dynamics of the total population are then projected forward in time, such that

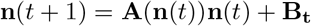

where **B**_**t**_ is a vector representing the number of births at time *t*, defined as,

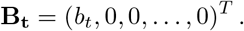

## Supplementary figures

**Figure S1:**
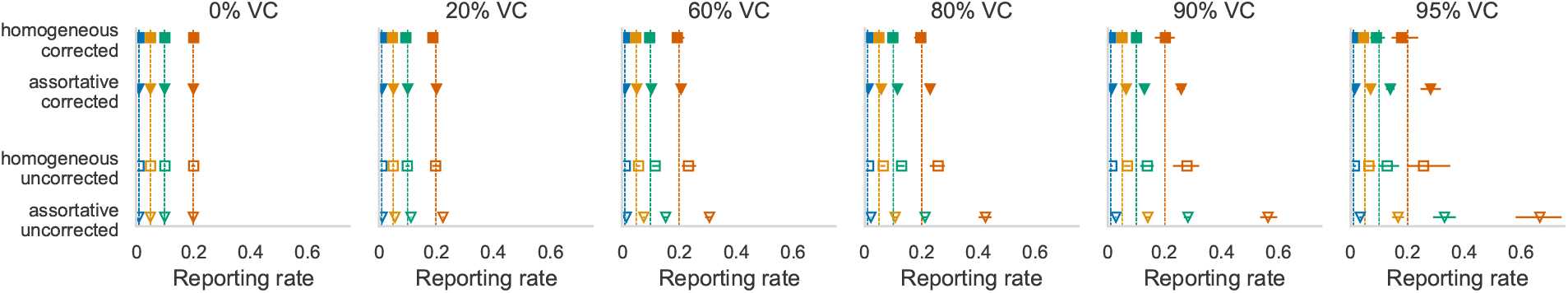
Estimated reporting fractions under homogeneous and assortative (Kenya-like) contact mixing consistent for different vaccination coverage (VC) and a high birth rate (40 births per thousand). Colors indicate different reporting fractions; vertical lines indicate simulated truth, and symbols indicate mean and range (horizontal lines) of estimates from 20 simulations.

**Figure S2:**
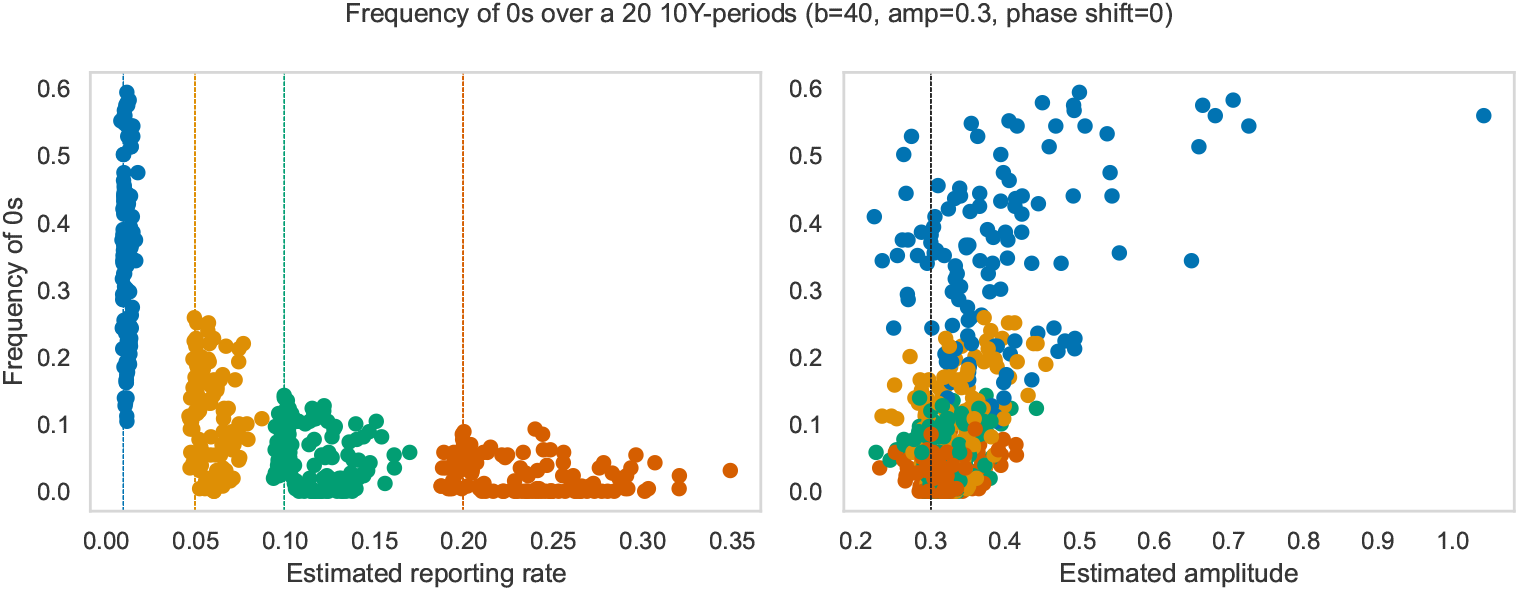
Frequency of zeroes in the reconstructed time series of incidence for different estimated reporting fraction and estimated amplitude for 20 ten-year time series. True reporting fractions are 0.01 (blue), 0.05 (yellow), 0.10 (green), and 0.20 (red). Here the true amplitude is 0.3 with 0 phase shift.

## References

[1] Anderson R, May R. Infectious Diseases of Humans: Dynamics and Control. New York, NY: Oxford University Press; 1991.

[2] Finkenstadt BF, Grenfell BT. Time series modelling of childhood diseases: a dynamical systems approach. Journal of the Royal Statistical Society, Series C. 2000;49:187–205.

[3] Keeling M, Rohani P. Modeling Infectious Diseases in Humans and Animals. Princeton, NJ: Princeton University Press; 2008.

[4] Bjørnstad ON, Finkenstädt BF, Grenfell BT. Dynamics of measles epidemics: Estimating scaling of transmission rates using a time series SIR Model. Ecological Society of America. 2002;72(2):169–84.

[5] Metcalf CJE, Bjørnstad ON, Grenfell BT, Andreasen V. Seasonality and comparative dynamics of six childhood infections in pre-vaccination Copenhagen. Proceedings of the Royal Society B: Biological Sciences. 2009 Sep;276(1676):4111–8. Available from: 10.1098/rspb.2009.1058.

[6] Gunning CE, Erhardt E, Wearing HJ. Conserved patterns of incomplete reporting in prevaccine era childhood diseases. Proceedings of the Royal Society B: Biological Sciences. 2014 Nov;281(1794):20140886. Available from: https://www.ncbi.nlm.nih.gov/pmc/articles/PMC4211441/.

[7] Edmunds WJ, Gay NJ, Kretzschmar M, Pebody RG, Wachmann H, ESEN Project European Seroepidemiology Network. The pre-vaccination epidemiology of measles, mumps and rubella in Europe: implications for modelling studies. Epidemiology and Infection. 2000 Dec;125(3):635–50. Edmunds2000.

[8] Ferrari MJ, Grais RF, Bharti N, Conlan AJK, Bjørnstad ON, Wolfson LJ, et al. The dynamics of measles in sub-Saharan Africa. Nature. 2008;451(7179):679–84.

[9] Becker AD, Wesolowski A, Bjørnstad ON, Grenfell BT. Long-term dynamics of measles in London: Titrating the impact of wars, the 1918 pandemic, and vaccination. PLoS Computational Biology. 2019 Sep;15(9):e1007305. Available from: https://www.ncbi.nlm.nih.gov/pmc/articles/PMC6742223/.

[10] Thakkar N, Gilani SSA, Hasan Q, McCarthy KA. Decreasing measles burden by optimizing campaign timing. Proceedings of the National Academy of Sciences of the United States of America. 2019;166(22):11069–73.

[11] Mahmud AS, Metcalf CJE, Grenfell BT. Comparative dynamics, seasonality in transmission, and predictability of childhood infections in Mexico. Epidemiology and Infection. 2017 Feb;145(3):607–25. Available from: https://www.ncbi.nlm.nih.gov/pmc/articles/PMC6020851/.

[12] World Health Organization. Measles. World Health Organization; 2023. Accessed on November 6, 2023. Available from: https://www.who.int/news-room/fact-sheets/detail/measles.

[13] GBD 2017 Population and Fertility Collaborators. Population and fertility by age and sex for 195 countries and territories, 1950–2017: a systematic analysis for the Global Burden of Disease Study 2017. The Lancet. 2018;392(10159):1995–2051. Available from: https://www.thelancet.com/journals/lancet/article/PIIS0140-6736(18)32278-5/fulltext.

[14] Fine PEM. Herd immunity: History, theory, practice. Epidemiologic Reviews. 1993;15(2):265–302. ISBN: 0193-936X (Print)\r0193-936X (Linking).

[15] Guerra FM, Bolotin S, Lim G, Heffernan J, Deeks SL, Li Y, et al. The basic reproduction number (R0) of measles: a systematic review. The Lancet Infectious Diseases. 2017;17(12):e420–8. Publisher: Elsevier Ltd. Available from: 10.1016/S1473-3099(17)30307-9.

[16] Prem K, Cook AR, Jit M. Projecting social contact matrices in 152 countries using contact surveys and demographic data. PLoS Computational Biology. 2017;13(9):e1005697.

[17] Gillespie DT. Approximate accelerated stochastic simulation of chemically reacting systems. The Journal of Chemical Physics. 2001 Jul;115(4):1716–33. Gillespie2001. Available from: 10.1063/1.1378322.

[18] United Nations, Department of Economic and Social Affairs. Population Division, World Population Prospects 2022 (Medium variant). United Nations; 2022. Online Edition.

[19] Central Intelligence Agency. Country comparisons – birth rate. Central Intelligence Agency; 2023. Accessed on November 6, 2023. Available from: https://www.cia.gov/the-world-factbook/field/birth-rate/country-comparison/.

[20] Liu WM, Levin SA, Iwasa Y. Influence of nonlinear incidence rates upon the behavior of SIRS epidemiological models. Journal of Mathematical Biology. 1986;23(2):187–204.

[21] Lau MSY, Becker AD, Korevaar HM, Caudron Q, Shaw DJ, Metcalf CJE, et al. A competing-risks model explains hierarchical spatial coupling of measles epidemics en route to national elimination. Nature Ecology & Evolution. 2020 Jul;4(7):934–9. Lau2020. Available from: https://www.nature.com/articles/s41559-020-1186-6.

[22] Klepac P, Caswell H. The stage-structured epidemic: linking disease and demography with a multi-state matrix approach model. Theoretical Ecology. 2011 Aug;4(3):301–19. Available from: 10.1007/s12080-010-0079-8.

[23] Klepac P, Pomeroy LW, Bjørnstad ON, Kuiken T, Osterhaus ADME, Rijks JM. Stage-structured transmission of phocine distemper virus in the Dutch 2002 outbreak. Proceedings of the Royal Society B: Biological Sciences. 2009 Jul;276(1666):2469–76. Available from: https://www.ncbi.nlm.nih.gov/pmc/articles/PMC2690464/.

[24] Metcalf CJE, Lessler J, Klepac P, Morice A, Grenfell BT, Bjørnstad ON. Structured models of infectious disease: Inference with discrete data. Theoretical Population Biology. 2012;82(4):275–82. ArXiv: NIHMS150003 Publisher: Elsevier Inc. ISBN: 1096-0325. Available from: 10.1016/j.tpb.2011.12.001.

[25] Winter AK, Martinez ME, Cutts FT, Moss WJ, Ferrari MJ, Mckee A, et al. Benefits and challenges in using seroprevalence data to inform models for measles and rubella elimination. Journal of Infectious Diseases. 2018;218(3):355–64.

